# Perceived acceptability of self-administered topical therapy for cervical precancer treatment among women undergoing cervical cancer screening in Kenya

**DOI:** 10.1101/2024.03.05.24303779

**Authors:** Chemtai Mungo, Grace K Ellis, Mercy Rop, Yating Zou, Jackton Omoto, Lisa Rahangdale

## Abstract

**Purpose:** Innovative strategies are urgently needed to meet the World Health Organization’s 2030 target of treating 90% of women with precancerous cervical lesions, especially in countries most affected by cervical cancer. We assessed the acceptability of self-administered intravaginal therapies for treating cervical precancer in women undergoing cervical cancer screening and precancer treatment in Kenya.

**Methods:** We conducted a cross-sectional study among women aged 18 to 65 years undergoing cervical cancer screening or precancer treatment between January and October 2023 in Kisumu County, Kenya. Participants completed a questionnaire about their perceptions and perceived acceptability of self- or provider-administered topical therapies for cervical precancer treatment. Quantitative data were summarized using descriptive statistics.

**Results:** A total of 379 questionnaires were completed. The median age of participants was 35 years (IQR 25-62), 62% had a primary education or less, and 71% earned $5 or less daily. All participants had been screened for cervical cancer, and 191 (51%) had received precancer treatment, primarily thermal ablation. Ninety-eight percent of participants were willing to use a self-administered intravaginal therapy for cervical precancer, if available. The majority, 91%, believed their male partner would support their use. Given a choice, 63% preferred self-admiration at home compared to provider-administration of a topical therapy in the clinic, citing time and cost savings. In multivariate analysis, married women were more likely to expect partner support for self-administration than single women. Participants preferred a therapy used less frequently but for a longer duration, compared to daily use therapy with a shorter duration of use.

**Conclusions:** Self-administered intravaginal therapies for cervical precancer treatment are highly acceptable among women undergoing screening and precancer treatment in Kenya.

## Introduction

Although cervical cancer is preventable, it is the second most common cancer among women worldwide.^1^ Global trends of cervical cancer represent a dire health inequity, with 85 percent of incident cases and 90 percent of deaths occurring in low- and middle-income countries (LMICs),^1^ due in part to lack of access to known primary and secondary prevention tools for girls and women in LMICs. In 2020, the World Health Organization (WHO) launched the 90/70/90 global strategy to eliminate cervical cancer, which calls for 90% HPV vaccination of girls, 70% of all women globally undergoing screening, and 90% of those diagnosed with cervical precancer or cancer adequately treated by 2030.^2^ Achieving these 90/70/90 targets would help reach the WHO elimination threshold of 4 or less cases of cervical cancer per 100,000 women, averting 62 million deaths in the next century.^3^ However, to achieve these targets, significant efforts are needed to close the cervical precancer treatment gaps among women in LMICs.

Current cervical precancer treatment methods include ablation or excision of precancerous lesions,^4^ both of which require specialized equipment and trained providers, making access to precancer treatment in LMICs a significant challenge,^5–10^ resulting in missed opportunities for secondary prevention and diminishing the public health impact of screening.^11^ There are high rates of loss-to-follow-up due to cost and transportation challenges when women screened in rural areas are referred to central facilities for treatment,^12^ as well as lack of adequate skilled healthcare providers to offer treatment.^8,10,13^ In a retrospective review of the 2011-2020 Kenya cervical cancer program data, linkage to treatment following positive screening results was 25-40%, even though a structured surveillance system was in place.^9^ This gap is consistently observed across multiple LMICs,^5,8,10,11,13^ and contributes to the disproportionate burden of cervical cancer. To meet the WHO’s 2030 target of treating 90% of women with cervical precancer globally, there remains an urgent need for practical and scalable strategies to close the precancer treatment gap in LMICs.

While no medical therapies are currently approved for cervical precancer treatment, the use of topical, non-excisional therapies for cervical precancer is an area of active investigation.^14–20^ The feasibility,^17,21,22^ acceptability,^15,23^ and efficacy of several topical therapies for cervical precancer treatment has been demonstrated by several studies in high-income countries (HICs),^17,23–25^ including randomized trials.^14,15,26–28^ One such drug is 5-Fluorouracil (5FU) cream.^14,15^ In a randomized U.S. trial of women with cervical intraepithelial neoplasia grade 2 (CIN2), participants were randomized to 6-month observation or self-administered intravaginal 5FU for primary treatment ^15^. Under intention-to-treat analysis, participants in the 5FU arm had a 1.62 relative risk of CIN2 disease regression (95% CI 1.10-2.56) compared to the observation arm (p=0.01), demonstrating the efficacy of self-administered 5FU cream for treating CIN2 disease. Similarly, in a 2020 U.S.-based Phase I proof-of-concept study among women with cervical intraepithelial neoplasia grade 2 or 3 (CIN2/3), primary treatment with self-administered intravaginal artesunate suppositories, which has been shown to have anti-HPV properties,^29–31^ was safe, well tolerated, and was associated with 67.9% CIN2/3 regression within 15 weeks.^17^ Both 5FU and artesunate are on the WHO List of essential medications,^32^ are generically available in LMICs, and could be repurposed as self-administered cervical precancer treatment in LMICs if backed by local feasibility, acceptability, and efficacy studies.

Self-administered topical therapies could be a scalable and cost-effective alternative to the less accessible provider-administered treatments in LMICs. Research on the acceptability on topical therapies for cervical precancer treatment in LMICs is needed to guide efficacy trials in these settings (Clinicaltrial.gov NCT05413811, NCT05362955, NCT06165614). To this end, we evaluated the perceived acceptability of topical therapies for cervical precancer treatment among women undergoing cervical cancer screening and precancer treatment in Kenya.

## Methods

### Study Design, Setting, and Recruitment

We conducted a cross-sectional study in Kisumu County, Kenya, between January and October 2023. Eligible participants were women aged 18 to 65 years who were undergoing cervical cancer screening or precancer treatment, primarily at outpatient HIV clinics. A convenience sampling technique was utilized where eligible participants were invited to participate and sequentially enrolled during the study period.

Kisumu County is one of 47 administrative units in Kenya,^40^ a country of 55.1 million in East Africa.^41^ Kisumu County is among the highest HIV burden regions in Kenya, with a 17.5% prevalence rate, compared to a national average prevalence of 4.9% in 2018.^42^ Cervical cancer is the leading cause of cancer death for women in Kenya, with an estimated 3,200 deaths in 2020.^11^

### Survey Development and Data Collection

The questionnaire collected sociodemographic as well as reproductive health information, including HIV status, cervical cancer screening, precancer treatment history, sources of health information, and history of intravaginal practices^33^ for medical or other reasons. The questionnaire assessed participants’ knowledge of HPV and cervical cancer risk factors and prevention methods. A script was used to explain self- or provider-administered intravaginal creams or suppositories for treating cervical precancer. Visual aids, including a pelvic model, sample vaginal suppositories, and applicators, were employed to enhance comprehension. Using the pelvic model, a trained research assistant demonstrated the use of an applicator to insert medication intravaginally and then a tampon to keep medication in place. Participants who had never used tampons examined sealed tampons. Other details provided included potential usage frequency (5FU once every other week for eight applications, artesunate daily for five days for three cycles), abstinence requirements (two to three days of abstinence after each 5FU application and none for artesunate), and the recommendation of consistent contraception use while using both therapies.

Participants were then asked about their perceptions of these topical therapies, willingness to use them, and preference of type. Participants were also asked about their preference for home self-administration versus provider administration in a health facility. They were asked whether they would be comfortable using tampons with these therapies and whether they believed their partner would support their use of topical therapies. Most questions were close-ended with the option to answer “yes,” “no”, or “unsure.” Participants selected from multiple choices to questions aimed at understanding the reasons behind their preferences. The questionnaire was based on WHO research toolkits^44,45^ and from studies used to evaluate the acceptability of health interventions in similar settings.^34^ We validated the questionnaire by having it reviewed by research assistants, practicing survey delivery in training sessions, and modification after the first ten participants. The questions on intravaginal practices were added midway through the study hence were not completed by all participants. The questionnaires were verbally administered in a private room by trained research assistants in the participant’s preferred language, either *English, Dholuo,* or *Swahili*. Each questionnaire took approximately 45 minutes, and participants were reimbursed 500 Kenya Shillings (approximately $5) for their time.

### Sample Size

No prior study has evaluated the acceptability of self-administered topical treatments for cervical precancer in LMICs. We defined acceptability as respondents answering yes to the question of their willingness to use a self-administered topical therapy for cervical precancer treatment. Assuming a conservative 60%-point estimate of acceptability at a 95% confidence interval and a ±5% error margin below which the intervention would not be ready for broader study. This is consistent with a recent study in Uganda evaluating the acceptability of integrated community-based HIV and cervical cancer screening,^34^ and a study on acceptability of HIV self-testing among key populations.^35^ Acceptability in these studies was defined as high (67%), moderate (34%-66%), or low (33%) based on population proportions. A power calculation estimated a minimum required sample size of 369 at a 95% confidence interval.

### Data Analysis

Data were collected via REDCap databases and analyzed with R version 4.1.0 (Vienna, Austria). Quantitative data were summarized with descriptive statistics, medians, and IQR, while qualitative data were shown as proportions. Due to a high yes response to acceptability, comparative analyses on acceptability predictors weren’t possible. Univariate logistic regression identified associations between clinical/demographic characteristics and preferences for self vs. provider-application, perceived partner support, and therapy type preferences based on treatment frequency and duration. ORs and 95% CIs were calculated using t-tests; F-tests provided p-values. Covariates significant in univariate analysis and other plausible ones were included in a multivariate logistic model to adjust ORs, 95% CIs, and p-values for predicting preferences (self-vs. provider-administration, partner support, therapy type).

### Ethical Approvals

The study received approval from Maseno University School of Medicine and the University of North Carolina, Chapel-Hill institutional review boards. All participants provided informed consent.

### Results

A total of 376 surveys were completed by women undergoing screening for cervical cancer. The median age of respondents was 35 years; 62% had primary school education or less (Table 1). The majority, 60%, were informally employed, and 71% reported a daily income of less than $5. Most participants were married or living with a partner (59%), and 58% were HIV-positive on self-report. All participants had previously been screened for cervical cancer, and 53% had a history of positive screening result, primarily following screening for HPV. Of the 200 participants with a history of positive screening results, 191 (96%) had received treatment, primarily thermal ablation. The majority of respondents had heard of cervical cancer (95%) and HPV (73%) previously (Supplementary Table 1).

**Table 1.** Sociodemographic, sexual, and reproductive health characteristics of women undergoing cervical cancer screening in western Kenya, n=376.

| **Characteristic** | **N (%)** |
| --- | --- |
| Age (median, range) | 35 [25, 62] |
| Age group, years |  |
| 25 – 34 | 184 (49) |
| 35 – 44 | 124 (33) |
| 45 or older | 68 (18) |
| Highest level of education |  |
| Less than primary | 85 (23) |
| Completed primary | 147 (39) |
| Completed secondary | 72 (19) |
| College or higher | 72 (19) |
| Occupation |  |
| Informal | 225 (60) |
| Formal | 59 (16) |
| Student | 11 (3) |
| None | 81 (21) |
| Marital status |  |
| Single/never married | 58 (15) |
| Married/living together | 221 (59) |
| Divorced/separated | 37 (10) |
| Widowed | 60 (16) |
| Number of children (median, range) | 3 [0, 10] |
| Daily income < 499 Kshs (\$5) | 266 (71) |
| < 100 Kshs (<\$1) | 47 (12) |
| 100–499 Kshs (\$1-4.99) | 219 (58) |
| 500–999 Kshs (\$5-10) | 73 (19) |
| > 1000 Kshs (>\$10) | 37 (10) |
| Electricity in home | 234 (62) |
| Tap water in home | 135 (36) |
| Christian religious affiliation | 374 (99) |
| History of smoking cigarettes (n=209) | 10 (3) |
| Currently smoking cigarettes | 2 (20) |
| Common source of health information<sup>1</sup> (n=503) |  |
| Health facility | 352 (70) |
| Radio | 91 (18) |
| Church | 19 (4) |
| Family/friends | 25 (5) |
| Other | 16 (3) |
| Gravidity (median, IQR) | 3 [2, 5] |
| Parity (median, IQR) | 3 [2, 4] |
| Age of first sexual intercourse (median, IQR) | 17.5 [16, 20] |
| Lifetime sexual partners (median, IQR) | 3 [2, 4] |
| History of STI on self-report | 60 (16) |
| Current use of contraception method | 212 (56) |
| Implant/ IUD | 110 (52) |
| Injectable | 44 (21) |
| Condoms | 21 (10) |
| OCP | 29 (14) |
| Tubal ligation | 8 (4) |
| HIV positive on self-report | 219 (58) |
| Currently on ARV | 218 (99) |
| On ARV treatment > 1 year | 217 (99) |
| Previously screened for cervical cancer | 376 (100) |
| Number of screening episodes (median, IQR) | 2 [1, 3] |
| Positive result on cervical cancer screening | 200 (53) |
| via HPV test | 167 (83) |
| via VIA test | 20 (10) |
| Negative or pending result | 176 (47) |
| Ever received cervical precancer treatment | 191 (51) |
| Cryotherapy | 7 (4) |
| Thermocoagulation | 167 (87) |
| LEEP | 3 (2) |
| Unknown | 14 (7) |

<sup>1</sup>Participants were able to select more than one response.

When asked about their perceptions of topical therapies, 98% of respondents would be willing to use a self-administered intravaginal treatment for cervical precancer and 88% believed their partner would supportive (Table 2). The vast majority (98%) would be willing to abstain from sex during topical treatment as necessary; 91% felt their male partner would be supportive of abstinence requirements. Similarly, the vast majority (89%) of women stated willingness to use dual contraception (hormonal and barrier) as part of topical treatment; 85% believed their partner would support use of dual contraception. Although few respondents knew what a tampon was (28%) or had used one previously (16%), following a brief description of what tampons were and their potential use as part of self-administered topical treatments, 92% stated their willingness to use one.

**Table 2.** Questions assessing perceptions and potential acceptability of self-administered treatment for cervical precancer among HIV-positive and HIV-negative women undergoing cervical cancer screening in western Kenya, n=376.

| **Question** | **Yes (%)** | **No (%)** | **Unsure (%)** |
| --- | --- | --- | --- |
| Willing to use self-administered intravaginal treatment at home | 370 (98) | 4 (1) | 2 (0.5) |
| Partner would support use of self-administered intravaginal treatment at home (n=370) | 327 (88) | 12 (3) | 31 (8) |
| Would have a private place at home to use self-administered treatment (n=72) | 72 (100) | 0 (0) | 0 (0) |
| Willing to abstain from sex for a certain period during treatment | 367 (98) | 4 (1) | 5 (1) |
| Partner would abstain from sex for a certain period during treatment | 342 (91) | 5 (1) | 29 (8) |
| Willing to use dual contraception during topical treatment | 333 (89) | 32 (8) | 11 (3) |
| Partner would support use of dual contraception during treatment | 319 (85) | 22 (6) | 35 (9) |
| Knows what a tampon is | 107 (28) | 258 (69) | 11 (3) |
| Has used a tampon | 59 (16) | 314 (83) | 3 (1) |
| Would be comfortable using a tampon as part of self-applied precancer treatment | 346 (92) | 20 (5) | 10 (3) |
| Partner would support use of tampon as part of self-applied precancer treatment | 327 (87) | 9 (2) | 40 (11) |
| Use of tampon as part of self-applied treatment would prevent acceptance of treatment | 34 (9) | 332 (88) | 10 (3) |

When asked about their preference for treatment location, 63% preferred self-administration at home, 32% preferred provider-administration in a facility, and 5% had no preference (Table 3). Reasons for preferring self-application at home included saving time (52%), and lower costs (45%). Reasons for preferring provider-application at clinic were perceptions of increased safety (56%) and uncertainty of correct self-application at home (43%). When asked their preference for 5FU or artesunate based on treatment duration (5FU once every other week for eight applications, artesunate daily for five days for three cycles), 64% preferred 5FU. This preference did not change when considering the abstinence requirements associated with 5FU use (Table 3).

**Table 3.** Preferences related to cervical precancer treatment with topical therapy among women undergoing cervical cancer screening in western Kenya, n=376.

| **Characteristic** | **N (%)** |
| --- | --- |
| **Preference for location of application** |  |
| Self-administration at home | 237 (63) |
| Provider-administration at clinic | 119 (32) |
| No preference or unsure | 20 (5) |
| Reason for self-administration at home preference<sup>1</sup> (n=258) |  |
| Saves time | 133 (52) |
| Cheaper/less transport | 115 (45) |
| Privacy | 5 (2) |
| Other | 5 (2) |
| Reason for provider-administration at clinic<sup>1</sup> (n=122) |  |
| Safer | 68 (56) |
| Unsure if can self-apply correctly | 52 (43) |
| Other | 2 (2) |
| **Preference for type of topical therapy** |  |
| Based on treatment duration |  |
| Preference for 5FU | 241 (64) |
| Preference for Artesunate | 135 (36) |
| Based on abstinence and contraception requirements |  |
| Preference for 5FU | 240 (64) |
| Preference for Artesunate | 136 (36) |

<sup>1</sup>Participants were able to select more than one response.

In multivariate analyses, women who were married or living together with their partner were 3.69 times more to expect partner support of use of self-administered therapies compared to single women (95% CI 1.47-9.26, p=0.007) (Table 4). Age, marital status, being HIV-positive and having heard of HPV before were associated with preference for self-compared to provider-administration of topical therapies on univariate analysis, although none were significant on multivariate analysis. Preference for 5FU versus artesunate based on frequency of application and treatment duration was independently predicted by participant’s education level, income, and having heard of HPV before (Table 4). Compared to those with less than a primary school education, participants who had completed a primary education were more likely to prefer 5FU over artesunate (AOR 2.23, 95% CI 1.23-4.03, p<0.001).

**Table 4.** Characteristics associated with perceived partner’s support of self-administered treatment, preference for self-application at home, and preference for type of topical therapy based on frequency of application and treatment duration use among women undergoing cervical cancer screening in western Kenya, n=376.

| Characteristic | n (%) |  | OR (95% CI) | p-value | Adjusted OR (95% CI) | p-value |
| --- | --- | --- | --- | --- | --- | --- |
|  | Partner support | Lack of partner support |  |  |  |  |
| Marital status |  |  |  |  |  |  |
| Single/never married | 46 (81) | 11 (19) | 1 (Reference) | 0.005 | 1 (Reference) | 0.007 |
| Married/living together | 203 (92) | 18 (8) | 2.70 (1.19, 6.11) |  | 3.69 (1.47, 9.26) |  |
| Divorced, separated, or widowed | 77 (79) | 20 (21) | 0.92 (0.40, 2.10) |  | 1.63 (0.59, 4.46) |  |
| Current contraception use |  |  |  |  |  |  |
| Yes | 191 (90) | 21 (10) | 1 (Reference) | 0.041 | 1 (Reference) | 0.297 |
| No | 135 (83) | 28 (17) | 0.53 (0.29, 0.97) |  | 0.70 (0.36, 1.36) |  |
|  | Preference for self-application | Preference for provider application |  |  |  |  |
| Age, years |  |  | 1.05 (1.02, 1.08) | 0.001 | 1.02 (0.98, 1.06) | 0.258 |
| Marital status |  |  |  |  |  |  |
| Single/never married | 26 (46) | 31 (54) | 1 (Reference) | 0.002 | 1 (Reference) | 0.070 |
| Married/living together | 72 (74) | 25 (26) | 1.98 (1.10, 3.58) |  | 1.86 (0.91, 3.80) |  |
| Divorced, separated, or widowed | 138 (62) | 83 (38) | 3.43 (1.72, 6.87) |  | 2.71 (1.16, 6.31) |  |
| Number of children |  |  | 1.13 (1.02, 1.24) | 0.018 | 1.05 (0.93, 1.20) | 0.424 |
| HIV-positive on self-report |  |  |  |  |  |  |
| Yes | 82 (54) | 69 (46) | 1 (Reference) | 0.003* | 1 (Reference) | 0.090* |
| No | 152 (69) | 67 (31) | 1.91 (1.24, 2.94) |  | 1.64 (0.93, 2.89) |  |
| Knows someone with cervical precancer or cancer |  |  |  |  |  |  |
| Yes | 125 (65) | 67 (35) | 1 (Reference) | 0.336 | 1 (Reference) | 0.029 |
| No | 111 (61) | 72 (39) | 0.81 (0.53, 1.24) |  | 0.59 (0.37, 0.95) |  |
| Heard of HPV before today |  |  |  |  |  |  |
| Yes | 187 (68) | 87 (32) | 1 (Reference) | <0.001 |  |  |
| No | 49 (49) | 52 (51) | 0.43 (0.27, 0.69) |  |  |  |
|  | Preference for 5-FU | Preference for Artesunate |  |  |  |  |
| Highest level of education |  |  |  |  |  |  |
| Less than primary school | 50 (59) | 35 (41) | 1 (Reference) | 0.005 | 1 (Reference) | <0.001 |
| Completed primary school | 81 (57) | 62 (43) | 2.01 (1.14, 3.55) |  | 2.23 (1.23, 4.03) |  |
| Completed secondary or higher | 109 (74) | 38 (26) | 0.91 (0.53, 1.58) |  | 0.74 (0.40, 1.37) |  |
| Daily income |  |  |  |  |  |  |
| < Kshs 500 | 162 (61) | 103 (39) | 1 (Reference) | 0.074 | 1 (Reference) | 0.011 |
| ≥ Kshs 500 | 78 (71) | 32 (29) | 1.55 (0.96, 2.51) |  | 1.98 (1.17, 3.36) |  |
| Heard of HPV before today |  |  |  |  |  |  |
| Yes | 185 (68) | 89 (32) | 1 (Reference) | 0.019 | 1 (Reference) | 0.005 |
| No | 55 (54) | 46 (46) | 0.57 (0.35, 0.91) |  | 0.48 (0.29, 0.80) |  |

## Discussion

To our knowledge, this is the first study to evaluate the perception and perceived acceptability of topical therapies for cervical precancer treatment among women undergoing cervical cancer screening and precancer treatment in a LMIC. We find strong support for topical therapies among surveyed women, nearly all of whom expressed a willingness to self-administer treatment, if available. Notably, half of the surveyed women had previously undergone excisional or ablative treatment for precancers. Additionally, most participants believed their male partners would support their use of self-administered topical treatments, including support of associated abstinence and contraception requirements. While most surveyed women had never used a tampon before, when educated about them, the majority felt comfortable with the idea of using tampons as part of topical treatment and did not perceive it as a barrier. When given the option of self-administration at home compared to provider-administration in a health facility, almost two-thirds of participants preferred self-administration, citing less cost, ease of access, and increased privacy. When participants were given a choice between two topical therapies, the majority favored topical 5FU over artesunate, despite the requirement to abstain from sex for a few days following 5FU use. In multivariate analysis, marital status was associated with higher perception of partner support of use of self-administered therapies.

Our findings suggest that the use of self-administered topical therapies is acceptable to women in LMICs and, if supported by local efficacy studies, may help bridge the notable gaps in cervical precancer treatment in these settings where the burden of cervical cancer is greatest. Current precancer treatments, which require trained healthcare providers, have limited reach due to the scarcity of professionals and difficulty accessing the services due to transport barriers, especially for women in rural areas without nearby referral centers. Our study and numerous others from LMICs have highlighted these access issues.^5,7,8,10,13^ In our study, the majority of participants pointed to lower transportation costs as a key reason for preferring self-administration of topical therapies over provider-administration in a health facility. This is further demonstrated by a qualitative study from Malawi where women with abnormal cervical cancer screening results cited lack of transportation and high associated costs as a major reason for not presenting for treatment.^8^ In this study, women who presented for treatment described the difficulty of travel, as many could not afford motorized transportation and were fatigued from journeying on foot. In contrast, self-administered topical therapies, if made available through rural pharmacies or dispensaries, could reach significantly more women. The use of self-administered therapies at home could also address other facility-level barriers to treatment, including lack of or non-functional treatment devices, which are frequently reported in LMICs.^10,12,36^ Similarly, self-applied therapies address issues of privacy, addressing concerns highlighted by our participants and echoed in other LMIC studies,^37^ where women identified the discomfort and invasiveness of speculum exams, particularly by male providers, as a barrier to seeking treatment.

Of note, while the majority preferred self-administration of topical therapies in our study, approximately a third of participants preferred provider-application, citing safety and doubts regarding their ability to correctly self-administer such therapies. This highlights the need for adequate education or appropriate patient selection if these therapies were made available, particularly in settings where women’s health literacy may be low. There are no current data that suggest increased safety or efficacy when topical therapies are applied by a health provider compared to self-application. We did, however, find that simple education including use of pictorials or models to explain pelvic anatomy can increase patient comfort with other unfamiliar components of topical treatment, such as tampon use. While few participants in our study knew what a tampon was nor had ever used one, following a brief explanation, most felt comfortable using them as part of treatment. This suggests that education can be crucial in alleviating women’s concerns related to safely and effectively self-applying topical intravaginal treatment. Further, our preliminary experience in an ongoing pilot clinical trial (NCT05362955) also supports evidence that women from Kenya with limited education and health literacy can safely use self-administered 5FU at home following adequate education and counseling.^38^

Another important finding from our study is the high perception of male partner support of women’s use of self-administered topical therapies, including support of abstinence and contraception use recommendations. While this requires exploration in studies of topical therapy use in LMICs, this is of significance as male partner support has been shown to impact the uptake of women’s reproductive health interventions, including cervical cancer prevention in sub-Saharan Africa.^39,40^ Male partner’s support of abstinence and contraception requirements associated with some topical therapies is especially crucial in settings where women may have reduced agency to negotiate this.^41^ In a recent qualitative study from Kenya, we report that with adequate education, men expressed support of their female partner’s use of topical therapies, including their abstinence requirements.^42^ More qualitative studies from different LMIC contexts are needed to inform this.

In this study, when offered options between two topical therapies for which early efficacy studies are available, most participants demonstrated a preference for 5-FU over artesunate, suggesting a preference for topical therapies used less frequently, even if associated with stricter abstinence requirements. Our assessment of the factors that impact women’s preferences for different tradeoffs was limited by the quantitative nature of our study. Such preferences can be explored further in qualitative studies or discrete choice experiments.

Our study has several limitations. Participants self-reported their preferences, and despite the use of trained research assistants who normalized all responses in the consenting process, it is possible that participants were influenced by social desirability bias and hence reported higher acceptability levels than would be observed under a different study design. Similarly, we surveyed women undergoing cervical cancer screening and oversampled women with a history of cervical precancer treatment as this intervention would be most applicable to them. It is possible that our findings may not be generalizable to women without a history of cervical cancer screening or those from settings different from the peri-urban area in Kenya where we recruited participants.

In summary, we report a high perceived acceptability of self-administered intravaginal therapy for cervical precancer treatment among women undergoing cervical cancer screening and precancer treatment in Kenya. Our findings demonstrate that self-administered topical therapies if backed by efficacy studies in LMICs, may play a crucial role in achieving the WHO’s 90% precancer treatment coverage globally and hence contribute towards eliminating this preventable cancer.

## Data Availability

Data is provided within the manuscript or supplementary information files and additional reasonable data requests can be accommodated.

---

**Supplementary Table 1.** Questions assessing knowledge of cervical cancer and HPV among women undergoing cervical cancer screening in western Kenya, n=376.

| **Question** | **Yes/True (%)** | **No/False (%)** | **Unsure (%)** |
| --- | --- | --- | --- |
| Heard of cervical cancer previously | 358 (95) | 18 (5) | 0 (0) |
| Heard that women can screen for cervical cancer | 366 (97) | 10 (3) | 0 (0) |
| Know anyone with cervical precancer or cancer | 192 (51) | 182 (48) | 2 (0.5) |
| Heard of human papillomavirus before today | 275 (73) | 89 (24) | 12 (3) |
| HPV can be transmitted via sexual intercourse | 222 (81) | 11 (4) | 42 (15) |
| Both men and women can be infected with HPV | 175 (64) | 56 (20) | 44 (16) |
| HPV infection is always symptomatic | 77 (28) | 136 (49) | 62 (22) |
| HPV can cause cervical cancer | 237 (86) | 16 (6) | 22 (8) |
| Having HIV increases a woman's risk of getting HPV or cervical cancer | 239 (87) | 16 (6) | 20 (7) |
| Smoking cigarettes increases risk of getting HPV or cervical cancer (n=209) | 108 (52) | 32 (15) | 69 (33) |
| Cervical cancer is preventable via vaccination or screening | 316 (84) | 23 (6) | 37 (10) |
| Only women with symptoms should get screened for cervical cancer | 78 (21) | 289 (77) | 9 (2) |
| Cervical cancer can be treated or cured if diagnosed early | 367 (98) | 5 (1) | 4 (1) |

## Notes

### Competing Interest Statement

The authors have declared no competing interest.

### Funding Statement

This research was supported by the by the Eunice Kennedy Shriver National Institute of Child Health & Human Development of the National Institutes of Health under Award Number K12HD103085 and the Victorias Secret Global Fund for Womens Cancers Career Development Award, in Partnership with Pelotonia Foundation and the American Association of Cancer Research (AACR). The content is solely the responsibility of the authors and does not necessarily represent the official views of the National Institutes of Health. The study funders have no role in the research.

### Author Declarations

The study received approval from Maseno University School of Medicine and the University of North Carolina, Chapel-Hill institutional review boards.

## References

1. Sung H, Ferlay J, Siegel RL, et al. Global Cancer Statistics 2020: GLOBOCAN Estimates of Incidence and Mortality Worldwide for 36 Cancers in 185 Countries. CA Cancer J Clin. 2021;71(3):209–249. doi:10.3322/caac.21660

2. WHO. A Global Strategy for elimination of cervical cancer. Accessed March 26, 2020. https://www.who.int/activities/a-global-strategy-for-elimination-of-cervical-cancer

3. Canfell K, Kim JJ, Brisson M, et al. Mortality impact of achieving WHO cervical cancer elimination targets: a comparative modelling analysis in 78 low-income and lower-middle-income countries. Lancet. 2020;395(10224):591–603. doi:10.1016/S0140-6736(20)30157-4

4. Castle PE, Murokora D, Perez C, Alvarez M, Quek SC, Campbell C. Treatment of cervical intraepithelial lesions. Int J Gynaecol Obstet. 2017;138 Suppl 1:20–25. doi:10.1002/IJGO.12191

5. Mungo C, Barker E, Randa M, Ambaka J, Osongo CO. Integration of cervical cancer screening into HIV/AIDS care in low-income countries: a moral imperative. Ecancermedicalscience. 2021;15:1237. doi:10.3332/ecancer.2021.1237

6. Mungo C, Ibrahim S, Bukusi EA, Truong HHM, Cohen CR, Huchko M. Scaling up cervical cancer prevention in Western Kenya: Treatment access following a community-based HPV testing approach. International Journal of Gynecology & Obstetrics. 2021;152(1):60–67. doi:10.1002/IJGO.13171

7. Beddoe AM. Elimination of cervical cancer: challenges for developing countries. Ecancermedicalscience. 2019;13. doi:10.3332/ECANCER.2019.975

8. Chapola J, Lee F, Bula A, et al. Barriers to follow-up after an abnormal cervical cancer screening result and the role of male partners: a qualitative study. BMJ Open. 2021;11(9):e049901. doi:10.1136/bmjopen-2021-049901

9. Mwenda V, Mburu W, Bor JP, et al. Cervical cancer programme, Kenya, 2011-2020: lessons to guide elimination as a public health problem. Ecancermedicalscience. 2022;16. doi:10.3332/ECANCER.2022.1442

10. Msyamboza KP, Phiri T, Sichali W, Kwenda W, Kachale F. Cervical cancer screening uptake and challenges in Malawi from 2011 to 2015: Retrospective cohort study. BMC Public Health. 2016;16(1):1–6. doi:10.1186/s12889-016-3530-y

11. Rohner E, Mulongo M, Pasipamire T, et al. Mapping the cervical cancer screening cascade among women living with HIV in Johannesburg, South Africaa. Int J Gynaecol Obstet. 2021;152(1):53–59. doi:10.1002/IJGO.13485

12. Khozaim K, Orang’O E, Christoffersen-Deb A, et al. Successes and challenges of establishing a cervical cancer screening and treatment program in western Kenya. Int J Gynaecol Obstet. 2014;124(1):12–18. doi:10.1016/J.IJGO.2013.06.035

13. Mungo, C Cohen, C, Bukusi, E Huchko M. Scaling up cervical cancer prevention in Western Kenya: Treatment access following a community-based HPV-testing approach. In: 33rd International Papillomavirus Conference (IPVC). ; 2020.

14. Maiman M, Watts DH, Andersen J, Clax P, Merino M, Kendall MA. Vaginal 5-fluorouracil for high-grade cervical dysplasia in human immunodeficiency virus infection: A randomized trial. Obstetrics and Gynecology. 1999;94(6):954–961. doi:10.1016/S0029-7844(99)00407-X

15. Rahangdale L, Lippmann QK, Garcia K, Budwit D, Smith JS, Le L Van. Topical 5-flourourcil for treamtent of cervical intraepithelial Neoplasia 2_J: a Randomized Controlled Trial. The American Journal of Obstetrics & Gynecology. 2014;210(4):314.e1-314.e8. doi:10.1016/j.ajog.2013.12.042

16. Desravines N, Hsu CH, Mohnot S, et al. Feasibility of 5-fluorouracil and imiquimod for the topical treatment of cervical intraepithelial neoplasias (CIN) 2/3. Int J Gynaecol Obstet. Published online July 2023. doi:10.1002/ijgo.14983

17. Trimble CL, Levinson K, Maldonado L, et al. A first-in-human proof-of-concept trial of intravaginal artesunate to treat cervical intraepithelial neoplasia 2/3 (CIN2/3). Gynecol Oncol. 2020;157(1):188–194. doi:10.1016/j.ygyno.2019.12.035

18. Fonseca BO, Possati-Resende JC, Salcedo MP, et al. Topical Imiquimod for the Treatment of High-Grade Squamous Intraepithelial Lesions of the Cervix: A Randomized Controlled Trial. Obstetrics and gynecology. 2021;137(6):1043–1053. doi:10.1097/AOG.0000000000004384

19. Lin CT, Qiu JT, Wang CJ, et al. Topical imiquimod treatment for human papillomavirus infection in patients with and without cervical/vaginal intraepithelial neoplasia. Taiwan J Obstet Gynecol. 2012;51(4):533–538. doi:10.1016/j.tjog.2012.09.006

20. De Witte CJ, Van De Sande AJM, Van Beekhuizen HJ, Koeneman MM, Kruse AJ, Gerestein CG. Imiquimod in cervical, vaginal and vulvar intraepithelial neoplasia: a review. Gynecol Oncol. 2015;139(2):377–384. doi:10.1016/J.YGYNO.2015.08.018

21. Desravines N, Hsu CH, Mohnot S, et al. Feasibility of 5-fluorouracil and imiquimod for the topical treatment of cervical intraepithelial neoplasias (CIN) 2/3. Int J Gynaecol Obstet. Published online 2023. doi:10.1002/IJGO.14983

22. Hampson L, Maranga IO, Masinde MS, et al. A Single-Arm, Proof-Of-Concept Trial of Lopimune (Lopinavir/Ritonavir) as a Treatment for HPV-Related Pre-Invasive Cervical Disease. PLoS One. 2016;11(1). doi:10.1371/JOURNAL.PONE.0147917

23. Desravines N, Miele K, Carlson R, Chibwesha C, Rahangdale L. Topical therapies for the treatment of cervical intraepithelial neoplasia (CIN) 2-3: A narrative review. Gynecol Oncol Rep. 2020;33:100608. doi:10.1016/j.gore.2020.100608

24. Hendriks N, Koeneman MM, van de Sande AJM, et al. Topical Imiquimod Treatment of High-grade Cervical Intraepithelial Neoplasia (TOPIC-3): A Nonrandomized Multicenter Study. J Immunother. 2022;45(3):180–186. doi:10.1097/CJI.0000000000000414

25. Desravines N, Chibwesha CJ, Rahangdale L. Low Dose 5-Fluorouracil Intravaginal Therapy for the Treatment of Cervical Intraepithelial Neoplasia 2/3: A Case Series. J Gynecol Surg. 2020;36(1):5–7. doi:10.1089/gyn.2019.0076

26. Grimm C, Polterauer S, Natter C, et al. Treatment of cervical intraepithelial neoplasia with topical imiquimod: A randomized controlled trial. Obstetrics and Gynecology. 2012;120(1):152–159. doi:10.1097/AOG.0b013e31825bc6e8

27. Polterauer S, Reich O, Widschwendter A, et al. Topical imiquimod compared with conization to treat cervical high-grade squamous intraepithelial lesions: Multicenter, randomized controlled trial. Gynecol Oncol. 2022;165(1):23–29. doi:10.1016/J.YGYNO.2022.01.033

28. Fonseca BO, Possati-Resende JC, Salcedo MP, et al. Topical Imiquimod for the Treatment of High-Grade Squamous Intraepithelial Lesions of the Cervix: A Randomized Controlled Trial. Obstetrics and Gynecology. 2021;137(6):1043–1053. doi:10.1097/AOG.0000000000004384

29. Disbrow GL, Baege AC, Kierpiec KA, et al. Dihydroartemisinin is cytotoxic to papillomavirus-expressing epithelial cells in vitro and in vivo. Cancer Res. 2005;65(23):10854–10861. doi:10.1158/0008-5472.CAN-05-1216

30. Efferth T, Dunstan H, Sauerbrey A, Miyachi H, Chitambar CR. The anti-malarial artesunate is also active against cancer. Int J Oncol. 2001;18(4):767–773. doi:10.3892/IJO.18.4.767

31. Augustin Y, Staines HM, Krishna S. Artemisinins as a novel anti-cancer therapy: Targeting a global cancer pandemic through drug repurposing. Pharmacol Ther. 2020;216. doi:10.1016/J.PHARMTHERA.2020.107706

32. World Health Organization (WHO). WHO Model List of Essential Medicines - 23rd List, 2023.; 2023. https://www.who.int/publications/i/item/WHO-MHP-HPS-EML-2023.02

33. Chisembele M, Rodriguez VJ, Brown MR, Jones DL, Alcaide ML. Intravaginal practices among young HIV-infected women in Lusaka, Zambia. Int J STD AIDS. 2018;29(2):164–171. doi:10.1177/0956462417721438

34. Mezei A, Trawin J, Payne B, et al. Acceptability of Integrated Community-Based HIV and Cervical Cancer Screening in Mayuge District, Uganda. JCO Glob Oncol. 2024;10(10). doi:10.1200/GO.22.00324

35. Figueroa C, Johnson C, Verster A, Baggaley R. Attitudes and Acceptability on HIV Self-testing Among Key Populations: A Literature Review. AIDS Behav. 2015;19(11):1949–1965. doi:10.1007/S10461-015-1097-8

36. Maza M, Figueroa R, Laskow B, et al. Effects of Maintenance on Quality of Performance of Cryotherapy Devices for Treatment of Precancerous Cervical Lesions. J Low Genit Tract Dis. 2018;22(1):47–51. doi:10.1097/LGT.0000000000000359

37. Knowledge and beliefs about cervical cancer screening among men in Kumasi, Ghana - PubMed. Accessed March 2, 2024. https://pubmed.ncbi.nlm.nih.gov/23661828/

38. Mungo C, Bukusi E, Kirkland GE, et al. Feasibility of adjuvant self-administered intravaginal 5-fluorouracil cream following primary treatment of cervical intraepithelial neoplasia grade 2 or 3 among women living with HIV in Kenya: study protocol for a pilot trial. medRxiv. Published online December 14, 2023. doi:10.1101/2023.12.13.23299916

39. Binka C, Doku DT, Nyarko SH, Awusabo-Asare K. Male support for cervical cancer screening and treatment in rural Ghana. PLoS One. 2019;14(11). doi:10.1371/JOURNAL.PONE.0224692

40. Adewumi K, Oketch SY, Choi Y, Huchko MJ. Female perspectives on male involvement in a human-papillomavirus-based cervical cancer-screening program in western Kenya. BMC Womens Health. 2019;19(1). doi:10.1186/S12905-019-0804-4

41. Seidu AA, Aboagye RG, Okyere J, et al. Women’s autonomy in household decision-making and safer sex negotiation in sub-Saharan Africa: An analysis of data from 27 Demographic and Health Surveys. SSM Popul Health. 2021;14. doi:10.1016/J.SSMPH.2021.100773

42. Mungo C, Adewumi K, Adoyo E, et al. “There is nothing that can prevent me from supporting her:” Men’s perspectives on their involvement and support of women’s use of topical therapy for cervical precancer treatment in Kenya. medRxiv. Published online December 26, 2023. doi:10.1101/2023.12.22.23300455

